# Uterine fluid transcriptome as potential non-invasive biomarker for predicting endometrial receptivity

**DOI:** 10.1101/2021.03.07.21253097

**Authors:** Aihua He, Hong Wu, Yangyun Zou, Cheng Wan, Jing Zhao, Qiong Zhang, Nenghui Liu, Donge Liu, Yumei Li, Jing Fu, Hui Li, Xi Huang, Tianli Yang, Chunxu Hu, Zhaojuan Hou, Yue Sun, Xin Dong, Jian Wu, Sijia Lu, Yanping Li

## Abstract

**Background:** The synchrony between the embryo and the receptive endometrium is essential for successful implantation. Therefore, a reliable non-invasive ER prediction method is highly demanded. We aimed to establish a method that could be used to predict endometrium receptivity non-invasively and to evaluate its clinical application potential in patients undergoing IVF.

**Methods:** The non-invasive RNA-seq based endometrial receptivity test (nirsERT) was established by sequencing and analyzing the RNA of uterine fluid from 48 IVF patients with normal ER. Subsequently, 22 IVF patients were recruited and analyzed the correlation between the predicted results of nirsERT and pregnancy outcomes.

**Results:** 87 marker genes and 3 hub genes were selected to establish the nirsERT. 10-fold cross-validation resulted in a mean accuracy of 93.0%. A small cohort retrospective observation showed that 77.8% (14/18) of IVF patients predicted with normal WOI had successful intrauterine pregnancies, while none of the 3 patients with displaced WOI had successful pregnancy.

**Conclusions:** nirsERT is potential for a non-invasive, accurate and same cycle testing for ER in reproductive clinic.

**Funding:** Funded by the National Natural Science Foundation of China (grant no. 8187061497) and the National Key Research and Developmental Program of China (grant no. 2018YFC1004800).

**Clinical trial number:** ChiCTR-DDD-17013375.

## Introduction

An ideal synchrony between the embryo and the receptive endometrium is necessary for successful implantation. The period of receptive endometrium, which referred to as window of implantation (WOI), normally occurs during the 19th to 24th day of a normal cycle. Previous studies demonstrated that the pregnancy rate would significantly reduce when implantation is not performed during the WOI [1, 2]. However, the optimal WOI lasts for less than 48 hours and varies wildly between individuals [3]. Abnormal endometrium receptivity (ER), including WOI shift and pathologic injury, has been observed in numerous patients with repeated implantation failure (RIF) [4–6]. Therefore, an approach of evaluating ER status is in urgent need, especially in the field of assisted reproductive technology (ART).

To fulfill this requirement, several methods had been proposed in the past decades, such as ultrasound examination [7–9], histologic analysis [10], and morphological markers [11–13]. But none had been proven to be an ideal predictor of endometrial receptivity. With the advance in molecular biological technologies, our understanding of molecular mechanism of embryo implantation has been significantly improved. In 2011, a 238 gene endometrial receptivity array (ERA) using RNA expression microarray was published by Diaz-Gimeno et al [14]. The ERA method is capable of identifying different stages of endometrial cycle, which are known as pre-receptive (PR), receptive (RE), and post-receptive (PO). The accuracy and reproducibility was proven to be reliable in subsequent studies [15–17]. Several studies have demonstrated that pregnancy outcomes of patients with RIF and infertile couples with conventional IVF [17, 18] can be improved by personalized embryo transfer (pET) guided by the ERA test. In addition, relevant results indicate that transcriptomic and proteomic markers provide promising approaches for ER assessment. Although numerous differentially expressed genes (DEGs) that are involved in endometrial receptivity have been revealed by previous studies, the overlap between these results is rather poor. One explanation might be that the sample size, individual differences and microarray platforms differ between studies. The next-generation, high-throughput RNA sequencing (RNA-seq) provides another powerful tool for analyzing the whole transcriptome comprehensively. RNA-seq is better than microarray at dynamic range, background noises, and identifying different transcripts [19, 20]. Another limitation for current diagnostic tools of endometrial receptivity has been the necessity of invasive tissue sampling by endometrial biopsy. The endometrial RNA expression profile could be altered due to the small injuries caused by invasive sampling [21]. Besides, local injury to the endometrium was reported to have a negative impact on implantation [22], therefore, it is inappropriate to perform endometrial tissue sampling test and guide implantation in a same active cycle. It is necessary to develop a non-invasive diagnostic tool to accurately predict WOI.

Uterine fluids is the important medium of communication between embryo and endometrium. It is an admixture of endometrial secretions, plasma transudates, and oviductal fluid [23]. Uterine fluid contains extracellular vesicles, RNAs, DNAs, regulatory proteins, ions, lipids and other bioactive factors and plays an important role in embryo implantation [24]. Thus, high throughput sequencing of uterine fluid provides an opportunity to find non-invasive biomarkers of endometrial receptivity for clinical use. Aspiration of uterine fluid prior to embryo transfer does not affect embryo implantation rate [25] also supports the feasibility of developing a non-invasive diagnostic tool based on uterine fluid. However, there are few transcriptional studies related to endometrial receptive markers from uterine fluid. A previous study [26] has identified a 53 candidate genes predictive of endometrial receptivity by using microarray technology to analyze uterine fluid, but it has not been developed into clinical diagnostic test.

Here, the aim of our study was to investigate the feasibility of predicting ER with biomarkers from uterine fluid, and to establish a non-invasive RNA-seq based endometrium receptivity test (nirsERT) which has the potential to be used in reproductive clinic.

## Methods

### Study Design

The main objective of this study was to establish a prediction tool for endometrial receptivity using transcriptome sequencing data, and to evaluate the feasibility of non-invasive endometrial receptivity test using uterine fluid specimen. Firstly, from November 2017 to December 2018, participants were recruited to identify differentially expressed genes (DEGs) among pre-receptive, receptive and post-receptive endometrium by transcriptome sequencing and expression profile analysis and to build the nirsERT model appling machine learning algorithm of random forest (RF). To limit interference from confounding variables affecting ER, the inclusion criteria for IVF patients were as follows: 20-39 years of age; body mass index (BMI)=18–25 kg/m2; secondary infertility with a history of a intrauterine pregnancy/pregnancies and undergoing the first IVF cycle due to tubal factors; primary infertility undergoing the first IVF cycle due to male factors; a regular menstrual cycle length (25-35 days) with spontaneous ovulation; normal ovarian reserve (baseline FSH < 10 mIU/mL, antimullerian hormone > 1.5 ng/ml, antral follicle count > 5); able to be followed up to assess the pregnancy outcome, and successful intrauterine pregnancy after the first embryo transfer (ET). The intrauterine pregnancy was defined as the presence of a gestational sac with or without fetal heart activity in the uterine cavity as evaluated by ultrasound 4–5 weeks after ET. To establish the prediction tool, normal ER status was defined with successful intrauterine pregnancy.

Secondly, from January to April 2019, participants were recruited to demonstrate the accuracy of nirsERT in predicting WOI. The inclusion criteria for patients who collected uterine fluid on the day of blastocysts transfer were as follows: 20-39 years of age; BMI = 18–25 kg/m^2^; ultrasound showed endometrial thickness *≥* 8 cm and endogenous serum progesterone level ≤1.2ng/ml on the day of progesterone administration/LH peak; the transferred embryos were high-quality blastocysts (blastocysts ≥ 3 BB on Day 5 and Day 6, graded based on the Gardner system) [27].

The following exclusion criteria were applied: endometrial diseases (including intrauterine adhesions, endometrial polyps, endometritis, endometrial tuberculosis, endometrial hyperplasia, and a thin endometrium); hydrosalpinx without proximal tubal ligation; submucous myomas, intramural hysteromyomas, or adenomyomas protruding towards the uterine cavity; endometriosis (stages III–IV); uterine malformations; and other medical or surgical co-morbidities were identified by consulting medical records, physical examination, blood test, B-ultrasound and X-ray examination.

In the validation group, all patients were performed nirsERT and were followed up to 4-5 weeks after ET to determine intrauterine pregnancy by ultrasound.

### Ethics statement

The present study was conducted at the Center for Reproductive Medicine at Xiangya Hospital of Central South University with permission by the Ethics Committee of Reproductive Medicine. This study was registered with the Chinese Clinical Trial Registry (No. ChiCTR-DDD-17013375).

### Uterine fluid collection, processing and transcriptome sequencing

All patients signed the written informed consent before sample collection. For patients included in the model construction, ultrasound was initiated from day 10 of the menstrual cycle preceding the IVF cycle to monitor ovulation. Blood LH levels were dynamically measured daily when the follicle diameter was > 14 mm. Patients continue to undergo daily ultrasound monitoring of ovulation until follicular discharge. Uterine fluid were respectively collected using embryo transfer catheter (Cook Medical; America) on days 5, 7, and 9 (LH+5, LH+7, and LH+9, respectively) after the LH surge (denoted as LH+0). For patients in the model validation group, the uterine fluid was collected on the day of blastocyst transfer before embryo transfer. (Transfers of frozen-thawed blastocysts were performed on the 7 days after the LH surge of natural cycle / the 5 days after progesterone supplementation of hormone replacement (HRT) cycles).

The sampling was performed as follows. The cervix was cleansed with saline before sampling. After the outer catheter of the embryo transfer catheter was inserted through the cervix to a depth of 4 cm from the external cervical os, the inner catheter was introduced into the uterine cavity to a point 1–2 cm from the uterine fundus to avoid contamination with cervical mucus. A 2.5 mL syringe was connected to the inner catheter and suction was applied. Inner catheter was withdrawn within the external catheter before external catheter was withdrawn from the uterus. Approximately 5-10uL of uterine fluid obtained were immediately placed into 20 uL of RNA-later buffer (AM7020; Thermo Fisher Scientific, Waltham, MA, USA) for RNA stabilization, sealed, and cryopreserved at −20°C. Sequencing analysis was carried out within 7 days after sampling.

Total RNA was extracted by using RNeasy Micro Kit (74004; Qiagen, city, state, country) according to the manufacturer’s instruction. Quality control of RNA was performed with Qubit HS RNA Kit (Q32855; Invitrogen) and Agilent Bioanalyzer 2100 (Agilent Technologies, city, state, country). Reverse transcription and library preparation were conducted using the MALBAC^®^ Platinum single cell RNA amplification kit and Transposon library Prep kit (KT110700796, and XY045, Yikon Genomics, Suzhou, China). Qualified libraries were sequenced by using the Illumina HiSeq 2500 platform with single-end reads length of 140bp. An average number of 5 million reads was generated for each library.

### Detection of differentially expressed genes

Differentially expressed genes (DEGs) among different endometrial receptivity conditions were identified by analysis of variance (ANOVA). The equation is stated as follows:

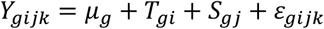

where *μ*_*g*_ represents the mean expression level of gene g; *T*_*gi*_ is gene-specific treatment effect referring to the status of being natural cycle or a hormone replacement therapy when uterine fluid was obtained, 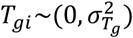; *S*_*gj*_ is gene-specific endometrial receptivity stage effect with three levels (pre-receptivity, receptivity, and post-receptivity), 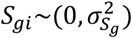 and *ε*_*gijk*_ is gene-dependent residual error, 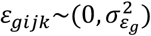. The F-test was applied to statistically assess the equality of variances between *S*_*j*_ and *ε*_*ijk*_ for each gene, showing whether the gene is differentially expressed among different endometrial receptivity stages. Because RNA-Seq analysis involves multiple statistical tests, the false discovery rate (FDR) was used to adjust the p-value (q-value) to provide statistical inference.

### Co-expression network construction and visualization

Co-expression modules in the endometrial receptivity process were detected by weighted gene co-expression network analysis (WGCAN) [28]. Applying WGCNA, we then identified key modules significantly correlated with endometrial receptivity stages. Cytoscape software was then used to visualize the interaction networks with different co-expression key modules [29].

### Biomarker identification and performance validation

To identify biomarkers for predictive model construction, post-hoc Tukey HSD (Honestly Significant Difference) test from ANOVA analysis was applied for pairwise comparisons of three receptive levels. Genes with significant differences of all pairwise test were detected for maximally distinguishing each receptive stage. Expression values of these biomarkers were then inputted as features for the machine learning method-random forest to train the pattern on three ER conditions (pre-receptivity, receptivity, and post-receptivity). The top important features (gene expression) were further selected by R package random Forest based on two measures (mean decrease accuracy and mean decrease gini). Out-of-bag (OOB) error, mean accuracy, sensitivity, specificity, positive predictive value, negative predictive value and F1 were determined from 10-fold cross-validation.

### Statistical analysis

Continuous data subject to a normal distribution were expressed as the mean ± SD. Continuous data subject to a skewed distribution were expressed as the median and inter-quartile range (IQR). Categorical data were expressed as counts and percentages, and were determined to be statistically significant using the chi-square test or Fisher’s exact test. A two-side P-value equal or less than 0.05 was considered to be statistically significant. Statistical analysis was performed using IBM SPSS software (Version 23.0, IBM Corp.)

## Results

### Participants

To establish the nirsERT model, we collected uterine fluid of three different receptive stages (pre-receptive, receptive and post-receptive) from infertile patients with normal WOI timing for RNA-seq. 69 participants were recruited and 21 patients who were not pregnant after the first embryo transfer were excluded, and 48 patients with successful intrauterine pregnancies were used to build nirsERT model (Figure 1). Baseline clinical characteristics are shown in supplementary Table S1.

**Figure. 1.**
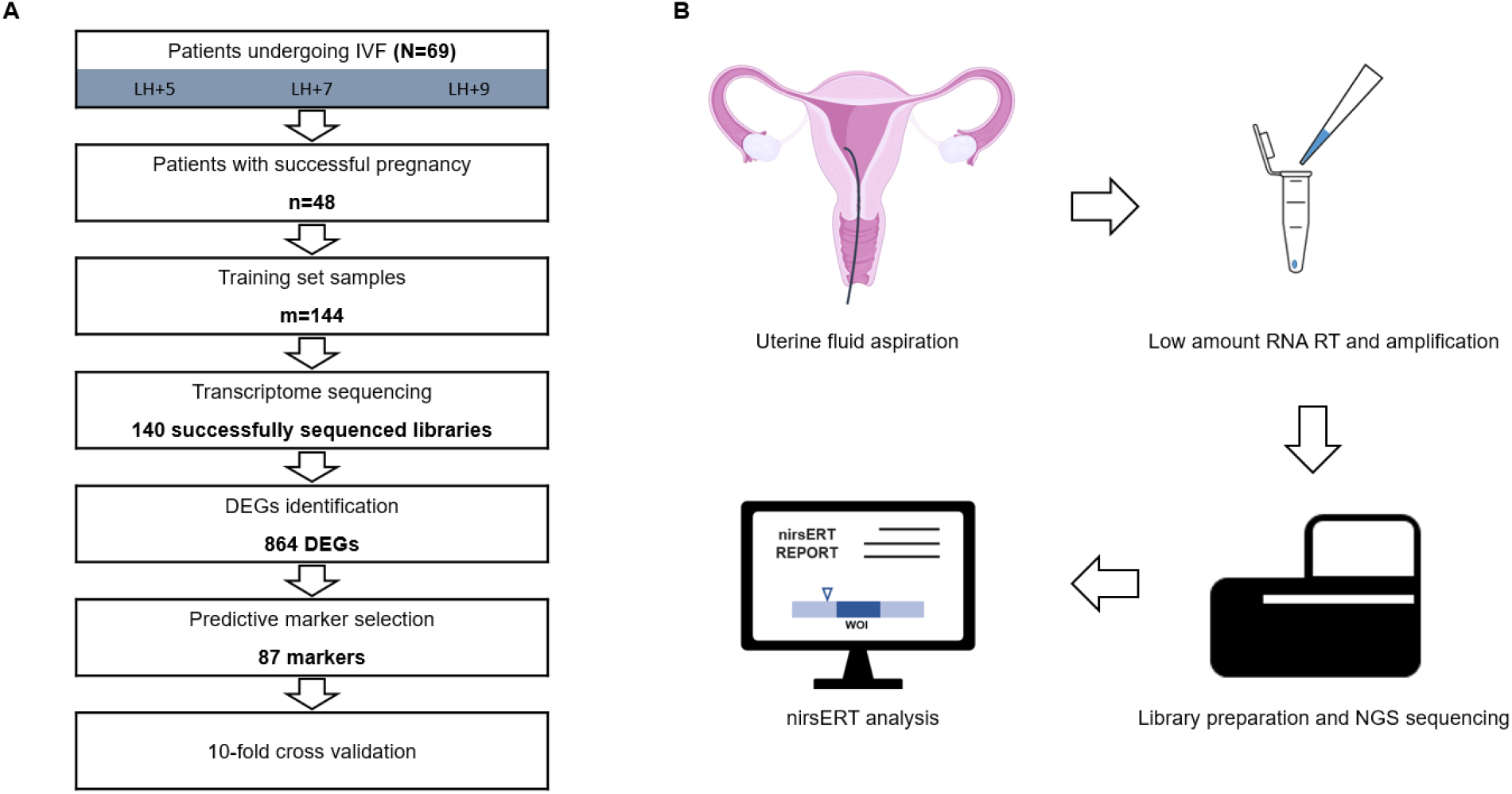
Flow diagram of Establishing and application of the non-invasive RNA-seq based endometrial receptivity test.

### Uterine fluid RNA extraction and sequencing

To perform the transcriptome sequencing, we collected 144 uterine fluid specimens from 48 participants and extracted total RNA by using commercial kit. As expected, the yield of RNA was relative low, ranging from 0 to 1160ng, with an average of 148ng. Almost one third of RNA samples were below detection limit of Qubit RNA HS assay kit (0.25ng/µL). Normally, it’s difficult to construct sequencing libraries starting with less than 1ng of total RNA. To address this, we utilized a commercial kit for reverse transcription and amplification with low amount of RNA.

We first validated the repeatability of transcriptome sequencing combined with above-mentioned kit (see supplementary methods). The Spearman correlation between different initial amounts of RNA was above 0.95, showing a high stability and repeatability of this method with at least 0.2ng RNA (Supplementary Figure S1). Then, we processed the 144 RNA samples according to the same protocol. As result, 140 NGS libraries were successfully constructed and sequenced, generating an average of 5.5 million raw reads per library. 632 million of high-quality reads, representing approximately 82.1% of raw data, were mapped to the human reference genome (Hg19). The number of mapped genes ranged from 9,591 to 17,913 in each library.

### DEGs detection and functional analysis

To identify differentially expressed genes (DEGs) among pre-receptivity, receptivity, and post-receptivity stages, ANOVA (analysis of variance) was applied to process the log2 transformed transcriptomic data. As result, 864 DEGs were detected within three different ER status. Notably, there are relatively more down-regulated DEGs between post-receptivity and receptivity status (Figure 2A). Unsupervised hierarchical clustering of the DEGs showed three distinct groups. GO analysis of these DEGs were conducted by DAVID tool (*20*). The DEGs were significantly enriched in 71 biological process (BP) terms, 38 cellular component (CC) terms and 25 molecular function (MF) terms. The top 1 enriched term for each category are signal transduction (GO:0007165), cytoplasm (GO:0005737), and protein binding (GO:0005515), respectively (Table 1 and Figure 2B).

**Table 1.** GO enrichment analysis of DEGs from uterine fluid samples.

| Category | Term | Gene count | p-value | Fold Enrichment | FDR |
| --- | --- | --- | --- | --- | --- |
| Biological Process | GO:0007165~signal transduction | 83 | 2.92E-05 | 1.59 | 0.05 |
|  | GO:0045944~positive regulation of transcription from RNA polymerase II promoter | 60 | 1.40E-02 | 1.36 | 22.49 |
|  | GO:0000122~negative regulation of transcription from RNA polymerase II promoter | 50 | 2.60E-03 | 1.54 | 4.61 |
|  | GO:0045893~positive regulation of transcription, DNA-templated | 39 | 1.98E-03 | 1.68 | 3.53 |
|  | GO:0006357~regulation of transcription from RNA polymerase II promoter | 36 | 8.56E-04 | 1.81 | 1.54 |
|  | GO:0006954~inflammatory response | 26 | 3.67E-02 | 1.52 | 49.24 |
|  | GO:0043065~positive regulation of apoptotic process | 24 | 9.19E-03 | 1.77 | 15.42 |
|  | GO:0050900~leukocyte migration | 21 | 5.05E-07 | 3.82 | 0.00 |
|  | GO:0001525~angiogenesis | 21 | 2.68E-03 | 2.09 | 4.75 |
|  | GO:0008360~regulation of cell shape | 18 | 1.87E-04 | 2.85 | 0.34 |
| Cellular Component | GO:0005737~cytoplasm | 298 | 7.41E-08 | 1.30 | 0.00 |
|  | GO:0005634~nucleus | 270 | 8.01E-03 | 1.13 | 10.94 |
|  | GO:0005829~cytosol | 200 | 8.78E-07 | 1.37 | 0.00 |
|  | GO:0070062~extracellular exosome | 183 | 1.55E-08 | 1.48 | 0.00 |
|  | GO:0005654~nucleoplasm | 157 | 5.89E-04 | 1.28 | 0.85 |
|  | GO:0016020~membrane | 146 | 2.40E-07 | 1.51 | 0.00 |
|  | GO:0005739~mitochondrion | 81 | 2.41E-03 | 1.38 | 3.42 |
|  | GO:0005615~extracellular space | 75 | 2.61E-02 | 1.27 | 31.66 |
|  | GO:0048471~perinuclear region of cytoplasm | 38 | 3.75E-02 | 1.39 | 42.34 |
|  | GO:0009986~cell surface | 34 | 3.70E-02 | 1.43 | 41.92 |
| Molecular Function | GO:0005515~protein binding | 492 | 2.66E-12 | 1.23 | 0.00 |
|  | GO:0044822~poly(A) RNA binding | 72 | 3.03E-03 | 1.41 | 4.64 |
|  | GO:0008270~zinc ion binding | 66 | 4.84E-02 | 1.24 | 54.05 |
|  | GO:0042803~protein homodimerization activity | 46 | 2.32E-02 | 1.39 | 30.79 |
|  | GO:0043565~sequence-specific DNA binding | 33 | 4.91E-02 | 1.40 | 54.63 |
|  | GO:0003682~chromatin binding | 32 | 1.88E-03 | 1.80 | 2.90 |
|  | GO:0005102~receptor binding | 27 | 1.07E-02 | 1.69 | 15.49 |
|  | GO:0003779~actin binding | 26 | 9.10E-04 | 2.06 | 1.42 |
|  | GO:0008134~transcription factor binding | 26 | 1.23E-03 | 2.02 | 1.90 |
|  | GO:0044212~transcription regulatory region DNA binding | 17 | 3.33E-02 | 1.76 | 41.17 |

**Figure. 2.**
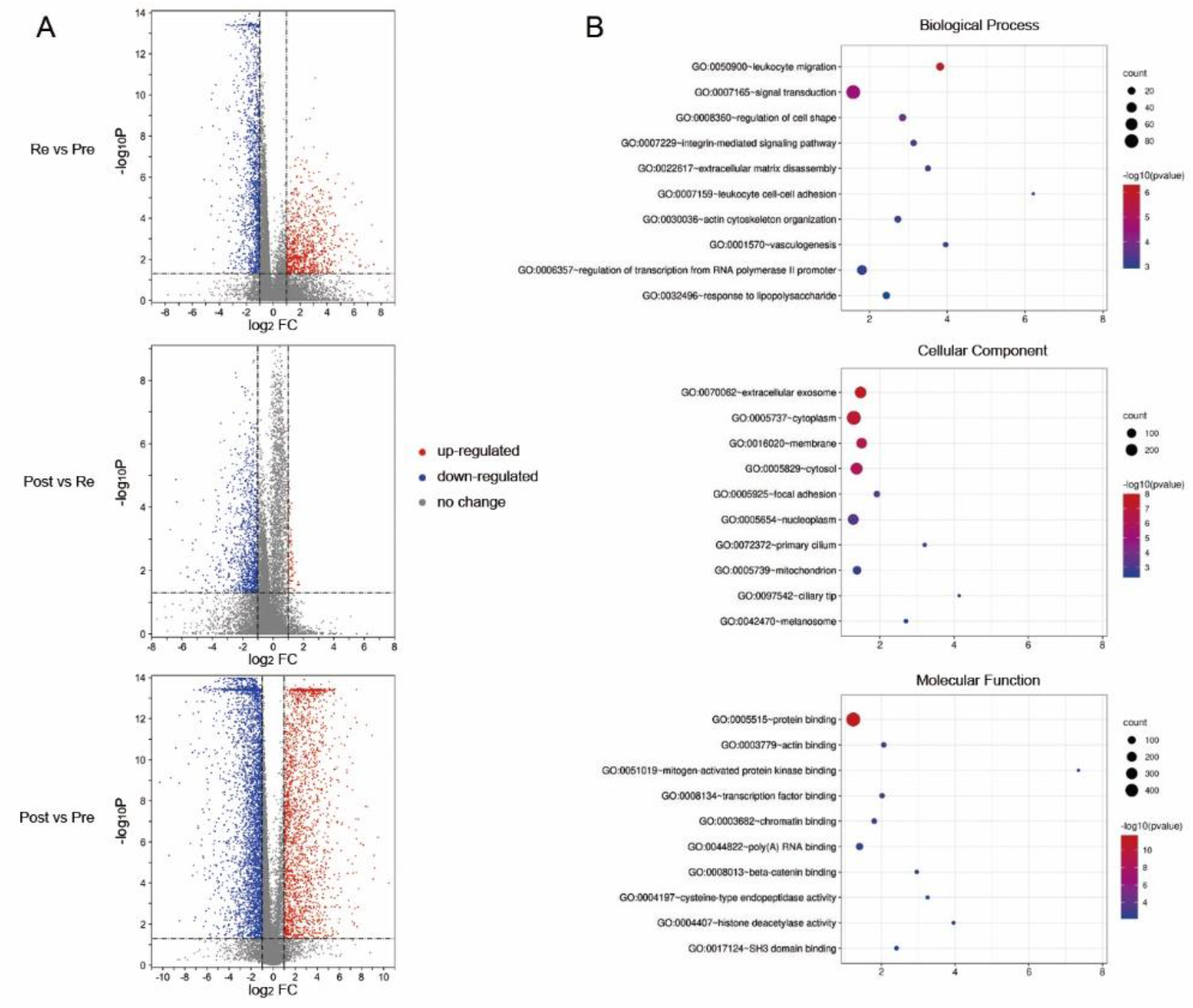
Differential expression analysis and functional enrichment among endometrial receptivity conditions.

To further investigate the functional module of DEGs in uterine fluid samples, we used the [weighted gene co-expression network analysis](https://www.ncbi.nlm.nih.gov/pmc/articles/PMC6380702/) (WGCNA) algorithm to analyze transcription regulatory networks. As result, 4 co-expression network modules with 3 being highly significant correlation with ER stages, which are MEturquoise, MEyellow and MEblue modules. Four hub genes ECI2 (MEturquoise), ATP6V1B2 (MEyellow), CXCL16 (MEblue) and SELP (MEgrey) were then identified based on the highest intramodular connectivity in four co-expression modules (Table 2). The MEturquoise module includes the most of DEGs, representing 59.1% (511/864) of total DEGs. It also shows the most significant correlation with ER stages with the correlation value of −0.7. Functional enrichment analysis shows genes in MEturquoise module involve in transcription regulation like epigenic modification related pathway; MEblue genes are enriched in GTPase mediated signal transduction, while MEyellow genes play roles in biomacromolecule transporting and cell-cell adherens junction. The result represents the whole involvement in endometrium-embryo crosstalk related biological processes of these DEGs detected in uterine fluid, which includes cell-cell communication, signal reception and transduction, and a series of cellular responses like transcription and translation of proteins responsible for embryo implantation.

**Table 2.** WGCNA analysis of DEGs from uterine fluid.

| Module | Number of genes | Hub gene | Module-receptivity relationships | DAVID cluster | *p-value | Enrichment score |
| --- | --- | --- | --- | --- | --- | --- |
| ME turquoise | 510 | ECI2 | -0.7 | GO:0016575~histone deacetylation | 0.0479 | 3.44 |
|  |  |  |  | GO:0004407~histone deacetylase activity | 0.0371 |  |
|  |  |  |  | GO:0016581~NuRD complex | 0.0416 |  |
| ME blue | 265 | CXCL16 | 0.55 | GO:0051056~regulation of small GTPase mediated signal transduction | 0.0192 | 3.5 |
|  |  |  |  | GO:0043547~positive regulation of GTPase activity | 0.0385 |  |
|  |  |  |  | GO:0005096~GTPase activator activity | 0.0557 |  |
| ME yellow | 78 | ATP6V1B2 | 0.69 | GO:0042470~melanosome | 0.0133 | 2.4 |
|  |  |  |  | GO:0045121~membrane raft | 0.0935 |  |
|  |  |  |  | GO:0005913~cell-cell adherens junction | 0.0935 |  |

\*: Benjamini adjusted p-value

### Establishing and validating the ER predictive tool

With Tukey test from ANOVA analysis, we selected genes with different expression in each pairwise comparisons of receptive stages (pre-receptivity versus receptivity, receptivity versus post-receptivity, and pre-receptivity versus post-receptivity). We therefore applied the expression pattern of these DEGs as training features for ER status classification using the random forest method. The random forest-based feature importance analysis with a top contribution to the model prediction by the mean decrease accuracy and Gini index was performed (*21*), resulting 87 predictive markers (Table 3). To strengthen the power of the predictive tool, we include three hub genes as additional markers (Figure 3), resulting the nirsERT. Linear discriminant analysis (LDA) showed three ER conditions (pre-receptivity, receptivity, and post-receptivity) were distinctly classified by the expression pattern of these transcriptomic markers (Figure 4A). To assess the performances of the present predictor, a 10-fold cross-validation was applied. We got mean accuracy of 93.0%, mean specificity of 95.9%, mean sensitivity of 90.0%. Uterine fluid samples of different ER conditions could be well separated by setting as a probability threshold of 0.6 (Figure 4B).

**Table 3.**
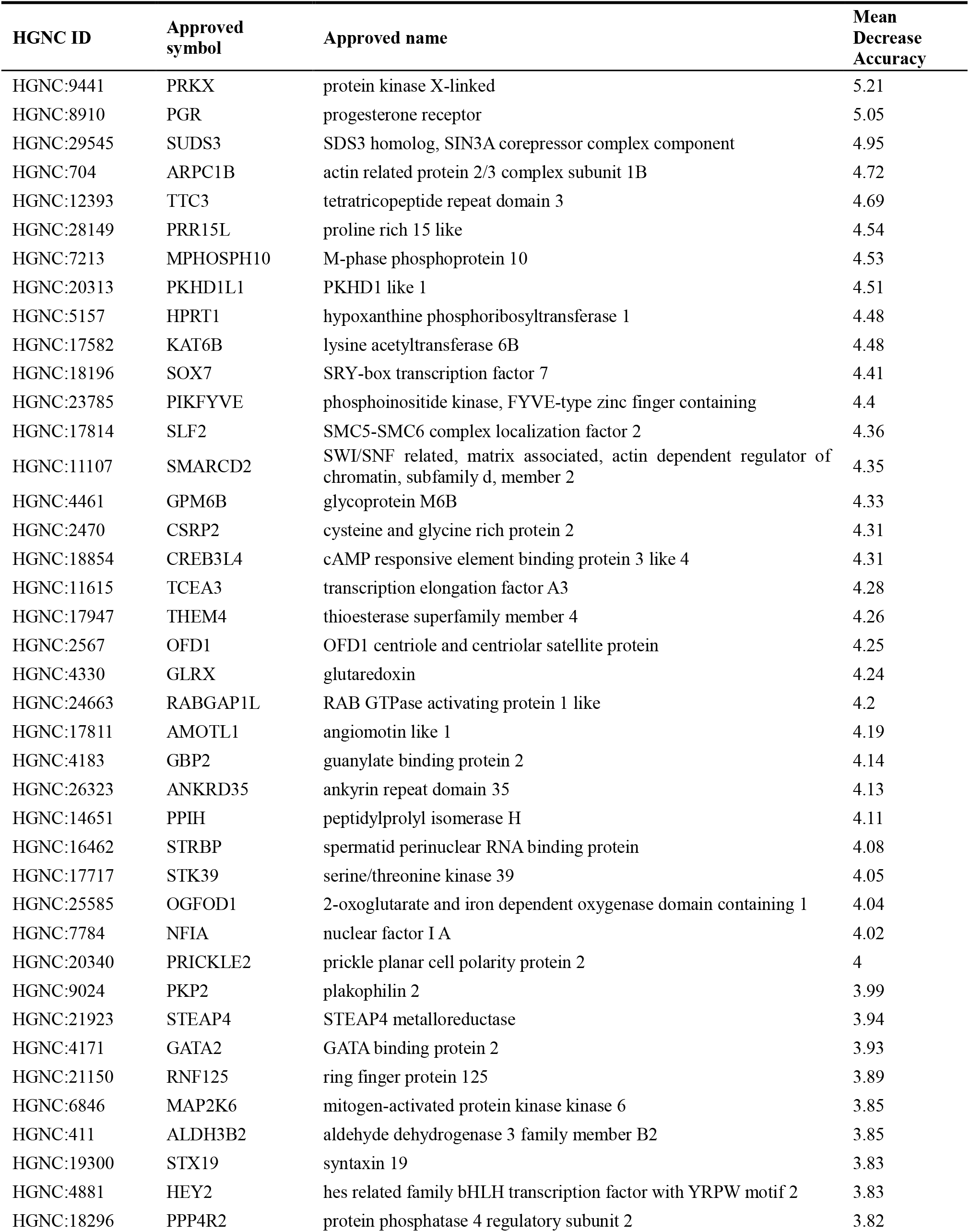
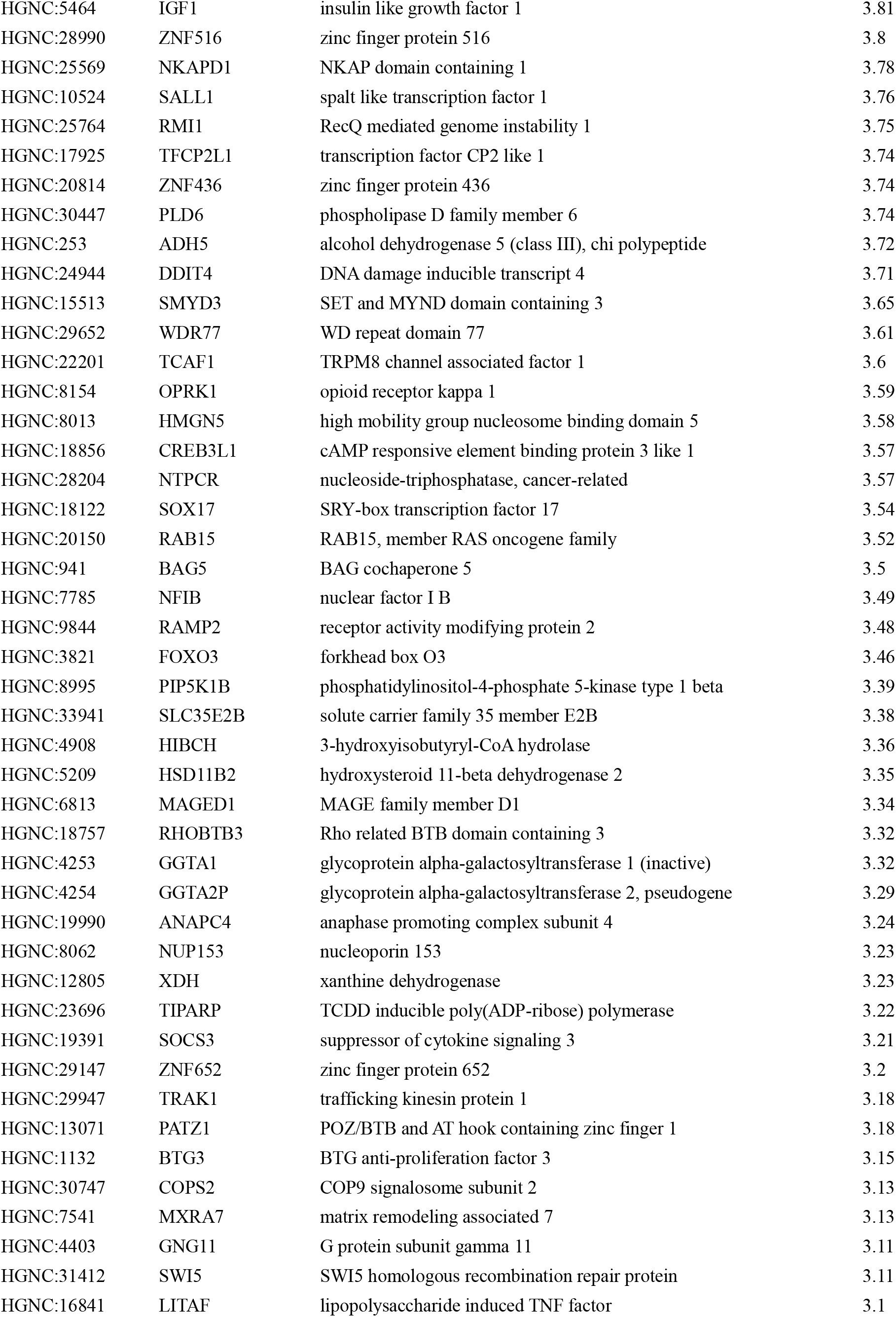
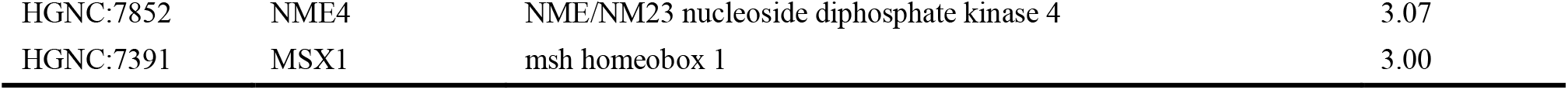
List of predictive markers selected by random forest algorithm

**Figure. 3.**
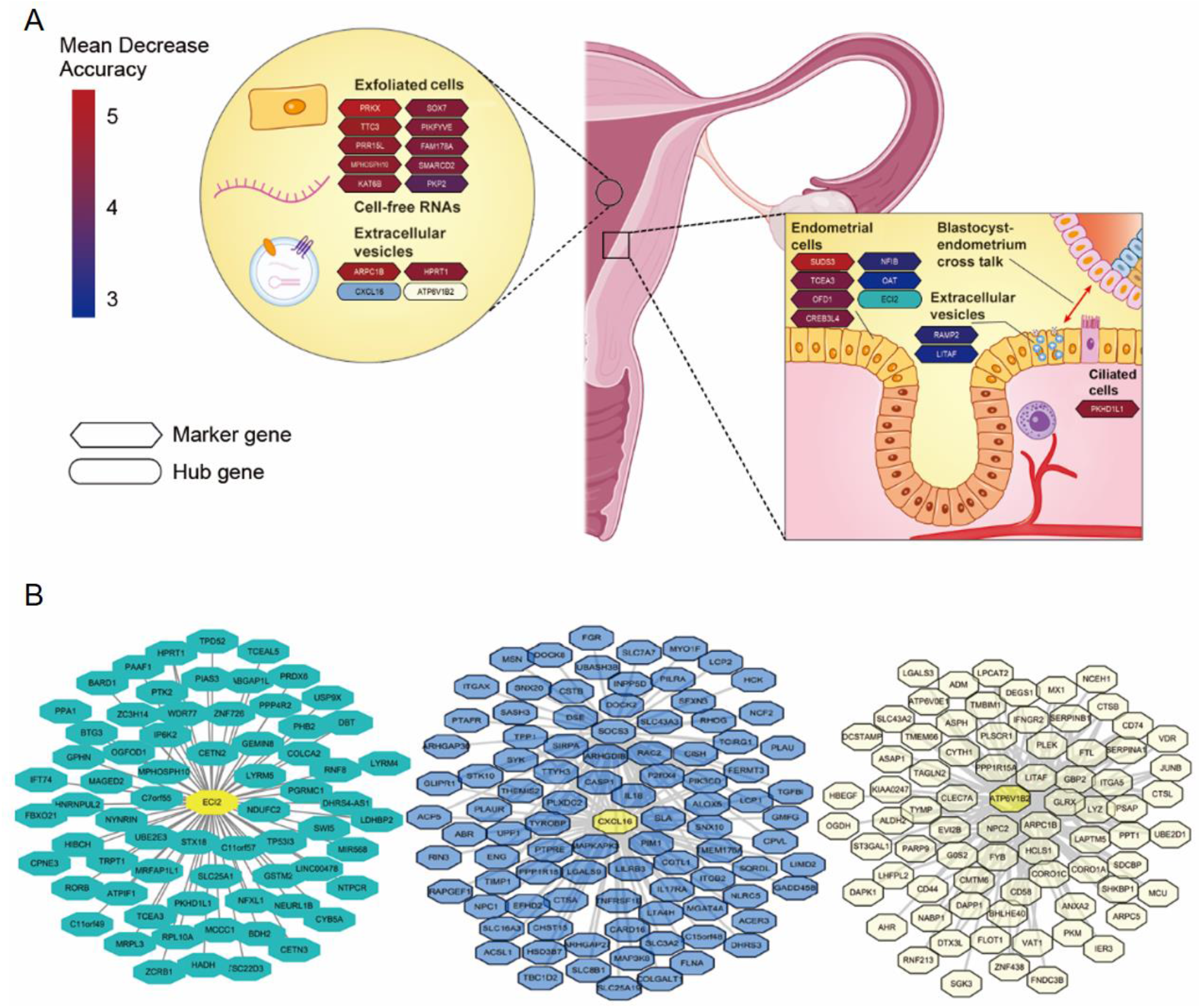
Partial predictive markers of nirsERT. A. Inferred source of marker and hub genes in nirsERT; B. Co-expression modules of uterine fluid DEGs generated with WGCNA.

**Figure. 4.**
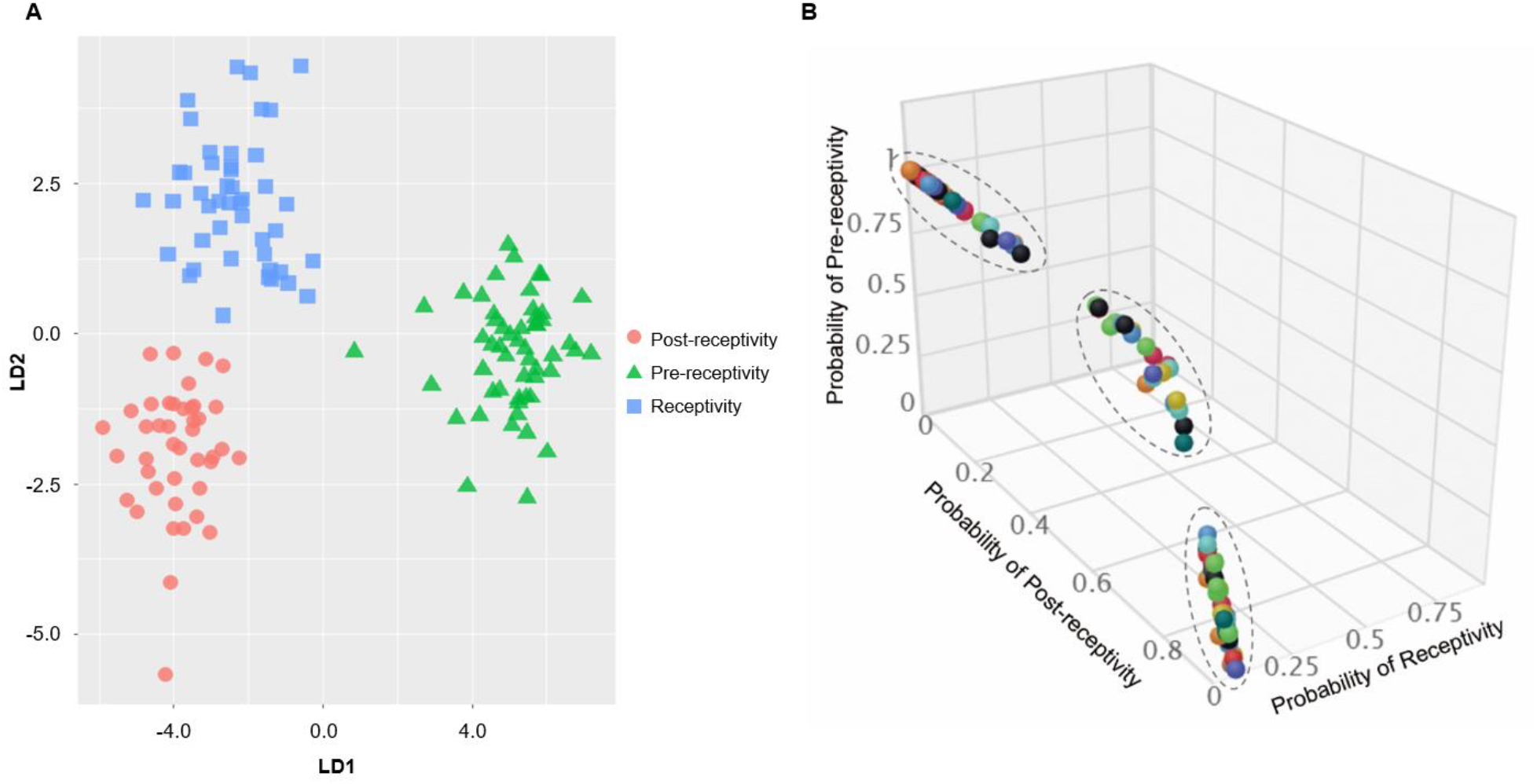
Establishment and validation of the nirsERT. A. Clustering the training set with LDA by using selected predictive markers; B. Prediction results of training set samples, with probability threshold of 0.6.

### Retrospective observation of a small cohort of patients undergoing IVF

To further evaluate the accuracy of the nirsERT, we analyzed the correlation between the predicted results of nirsERT and pregnancy outcomes. 22 uterine fluid samples from IVF patients were collected on the day of blastocyst transfer before embryo transfer and tested. The intrauterine pregnancy was determined by ultrasound 28 days after embryos transferred. The success rate of sequencing was 95.4% (21/22), with 1 libraries failed to pass the quality control procedure. As result, 18 patients (85.7%, 18/21) were predicted with normal WOI, whereas 3 (14.3%, 3/21) and 0 were predicted with delayed and advanced WOI, respectively. The intrauterine pregnancy rate (IPR) was 77.8% (14/18) among patients with normal WOI. There was no successful pregnancy in patients with displaced WOI, which was significantly different from those with normal WOI (*P*<0.05). The overall IPR in all patients was 63.6% (14/22) (Table 4).

**Table 4.** nirsERT results and clinical outcomes of 22 patients undergoing IVF.

|  | Normal WOI | Displaced WOI<br>delayed | WOI<br>advanced | P-value | Detection<br>failed | Total |
| --- | --- | --- | --- | --- | --- | --- |
| Date of transfer | LH+7/P+5 | LH+7/P+5 | LH+7/P+5 |  | LH+7/P+5 |  |
| Predicted result | Receptivity | Pre-receptivity | Post-receptivity |  | / |  |
| No. of patients | 18 | 3 | 0 |  | 1 | 22 |
| No. of intrauterine pregnancy | 14 | 0 | 0 |  | 0 | 14 |
| Intrauterine pregnancy rate | 77.8%(14/18) | 0 | 0 | 0.026 | 0 | 63.6%(14/22) |

## Discussion

In the past decades, researchers have investigated a variety of approaches to evaluate the condition of endometrial receptivity. However, limited progress had been made until the transcriptomic markers were established [26, 30]. Diagnostic tool result from endometrial tissue transcriptome is accurate and reproducible, but the application was also hindered by the necessity of invasive sampling. Thus, developing a non-invasive, precise and reliable method of ERT is one of the major challenges in reproductive medicine. In this study, a non-invasive ERT method based on RNA-seq was described for the first time, and it had the following benefits compared with previous studies: (1) RNA-seq could be used to identify more genes and in a more accurate manner than the conventional gene microarray; (2) Rather than two time points sampling, we collected samples of uterine fluid at three different time points, the pre-receptive, receptive, and post-receptive. Thus, the time span was shorten and a highly correlated sample cohort was established; (3) over 800 of DEGs in uterine fluid were analyzed, providing insight into function and role of multiple genes in the process of embryo implantation. It is difficult to perform transcriptome sequencing with uterine fluid samples, as nearly 1/3 of the samples yielded total RNA less than 0.25ng/µL. To address this, we utilized a commercial kit designed for single-cell RNA reverse transcription and amplification. The results showed a high stability and repeatability, the Spearman correlation between different amounts of total RNA ranging from 0.2ng to 20ng were above 0.98. By using this kit, we successfully prepared 140 RNA-seq libraries and constructed the training dataset. However, there were still 4 libraries failed to pass the quality control, we assume this might be caused by extremely low amount of RNA in these uterine fluid samples. To ensure the availability of nirsERT, it is important to investigate the distribution of the amount of total RNA in population. Besides, the improvement of uterine fluid aspiration could be helpful in further studies.

According to our previous study (preprinted) [31], there were 3571 DEGs identified from endometrial tissue among there ER status, a predictive tool (rsERT) consisted of 175 marker genes was established based on these DEGs. In current, a total of 864 DEGs were identified, including 468 common DEGs and 396 uterine fluid specific DEGs, compared with the study of rsERT. We found these common DEGs are significantly enriched in extracellular exosome (GO:0070062), cytoplasm (GO:0005737), cytosol (GO:0005829), nucleoplasm (GO:0005654) and protein binding (GO:0005515), which support the scenario that RNAs in uterine fluid originated from endometrial tissue cell with exosome secreted the outside of the cell. Unexpectedly, 396 DEGs were specifically observed in uterine fluid samples. These genes significantly involve in integrin-mediated signaling pathway (GO:0007229) and immune responses like leukocyte migration (GO:0050900), inflammatory response (GO:0006954) and response to lipopolysaccharide (GO:0032496). Besides, approximately 38.2% (330 of 864) of total DEGs were previously reported [13, 14, 32–35], while 61.8% (534 of 864) were first identified to be differently expressed in all three status of receptive. Our findings highlight the importance of genes involved in protein binding, signal transduction, and leukocyte migration in the uterine fluid. For instance, DEGs enriched in extracellular exosome (GO:0070062), including SLC25A1 (ENSG00000100075), PLSCR1 (ENSG00000188313), and NME3 (ENSG00000103024) were observed to be significantly related to the dynamic change of the sequential receptivity stages, which are assumed to mediate the communication between endometrium and embryo. Other cellular responses and signal transduction-related factors, e.g., RAC2 (ENSG00000128340) and ESR1 (ENSG00000091831), were also observed in our study (see Supplementary Table S2 and S3).

Four hub genes, ECI2, ATP6V1B2, CXCL16 and SELP were identified by using WGCNA analysis. ECI2 encodes a key mitochondrial enzyme involved in beta-oxidation of unsaturated fatty acids which may provide energy necessary for embryo implantation course. SELP implies the possible mechanism of P-selectin mediated cell adhesion in endometrium-embryo interaction. CXCL16 and its receptor CXCR6 were reported to play role in the decidualization during pregnancy [36]. ATP6V1B2 (ATPase H+ Transporting V1 Subunit B2) is a transmembrane transporter, which may be responsible for transporting biomacromolecule like secretory protein to its target location like extracellular matrix.

nirsERT consisting of 87 markers and 3 hub genes were selected by using random forest algorithm among 864 DEGs was established. We compared two predictive tools, nirsERT and rsERT established by using endometrial tissue samples in our previous study, only 22 markers were shared for both uterine fluid and tissue samples (Supplementary Table S4). According to the Human Protein Atlas, proteins generated by these gene locate in variety of subcellular locations [24], such as vesicle (BAG5, RAMP2), nucleus or nucleoplasm (ZNF652, TRAK1), cytosol (MAP2K6, RNF125) and cell junctions (PKP2). Besides, High correlation of expression pattern for these genes were observed between uterine fluid and endometrial tissue samples (Supplementary Figure S2). The results indicate the source of the common markers could be exfoliated endometrial cells or extracellular vesicles. The performance of nirsERT with rsERT was also compared study by using a same standard. 10-fold cross-validation resulted in comparable mean accuracy (93.0% vs 98.4%), mean specificity (95.9% vs 98.9%) and mean sensitivity (90% vs 97.8%).

We also investigated the selected markers in previous studies [14, 26], poor commonness was observed (Supplementary Figure S3). No common marker is selected in all three studies. There is no universal standard of selecting marker genes for endometrial receptivity, the mechanism of uterine transcriptomic changes during the process of embryo implantation is still unrevealed. Further investigations are required for raising power and reproducibility of the endometrial receptivity prediction.

To verify the accuracy of nirsERT in predicting endometrial receptivity, the uterine fluid collected on the day of blastocyst transfer was performed nirsERT. The accuracy of nirsERT prediction was evaluated by analyzing the correlation between the predicted results and subsequent pregnancy outcomes. The results showed that 77.8% (14/18) of patients predicted with normal WOI had successful intrauterine pregnancies, while none of the 3 patients with displaced WOI had successful pregnancy. It is suggested that the failure of embryo implantation in patients with displaced WOI may be the result of embryo-endometrial asynchrony. Although there are still four unsuccessful intrauterine pregnancies in patients with normal WOI predicted by nirsERT, 77.8% of IRP is consistent with the view that endometrial factors are responsible for about two-thirds of embryo implantation [37, 38]. Therefore, the results also further clinically validated the accuracy of nirsERT in predicting WOI. Personalized embryo transfer (pET) guided by nirsERT can possibly contribute to restore the synchronicity of embryonic and endometrial development which promoted successful embryo implantation. In addition, clinical pregnancy rate of routine blastocyst transplantation in our center was 55-60%, while the overall intrauterine pregnancy rate of patients with aspiration of uterine fluid on the day of embryo transfer was 63.6%, suggesting that aspiration of uterine fluid did not affect the embryo implantation. nirsERT has the potential to detect and guide pET in a same active cycle contributing to the successful embryo implantation.

It follows that our method provides currently the most promising approach for ideal pET. However, there is an issue has to confront, which is that whether nirsERT can improve the pregnancy outcomes of IVF patients by guiding pET has not been demonstrated yet, and we think it would be better to design a randomized clinical trial in the future to verify the clinical application value of nirsERT. In addition, the mechanism of endometrial receptivity marker genes also needs further investigation so as to provide theoretical basis for clinical treatment strategy.

## Conclusions

In conclusion, we established a non-invasive RNA-seq based endometrial receptivity test (nirsERT) by transcriptome sequencing analysis of uterine fluid combined with random forest algorithm. Endometrial receptive DEGs in uterine fluid may be derived from endometrial tissue cells and have an independent role in embryo implantation. nirsERT has the equivalent accuracy of endometrial receptive prediction to endometrium samples and is potential for a non-invasive, accurate and same cycle testing for endometrium receptivity in reproductive clinic.

## Supporting information

supplementary Table S1_clinical characteristics of participants

supplementary Table S2_functional enrichment of uterine fluid DEGs

supplementary Table S3_GO enrichment analysis of common and specific DEGs from uterine fluid samples

supplementary Table S4_common markers for nirsERT and rsERT

## Data Availability

All data related to this study can be obtained from the corresponding authors.

## Acknowledgments

We thank all patients and their family for the participation. We also thank Elsevier Author Services for preparation the Figure 3.

## Conflict of Interest

All the authors have read the manuscript and approved this for submission as well as no competing interests.

## Notes

### Competing Interest Statement

The authors have declared no competing interest.

### Clinical Trial

ChiCTR-DDD-17013375

