## supplementary methods and Figures for "Uterine fluid transcriptome as potential non-invasive biomarker for predicting endometrial receptivity"

**Materials and Methods**

**Validation of transcriptome sequencing with low amount of RNA**

To address the difficulty of constructing sequencing libraries starting with less than 1ng of total RNA, we utilized the MALBAC^®^ Platinum single cell RNA amplification kit (KT110700796, Yikon Genomics, Suzhou, China) for reverse transcription and amplification with low amount of RNA. To ensure the stability and repeatability of this kit, we first performed a pilot study with different amount of total RNA.

Total RNA was extracted from uterine fluid samples by using the RNeasy Micro Kit (74004; Qiagen, city, state, country) according to the manufacturer's instruction. Quality control of RNA was performed with Qubit HS RNA Kit (Q32855; Invitrogen) and Agilent Bioanalyzer 2100 (Agilent Technologies, city, state, country). Then, a set of RNA extracted from three sequential uterine fluid samples was selected, each with the yield over 200ng. We obtained RNA with the amount of 0.02ng, 0.2ng, 2ng, 20ng and 100ng by gradient dilution with RNase-free H_2_O. At last, these diluted RNA was processed with commercial kits to construct sequencing libraries and sequenced with the Illumina HiSeq 2500 platform. An average number of 5 million reads was generated for each library. Fragments per Kilobase Million (FPKM) was calculated by Cufflinks (*1*), the Spearman correlation was then calculated to compare the differences between different initial amounts of RNA.

**Figures**

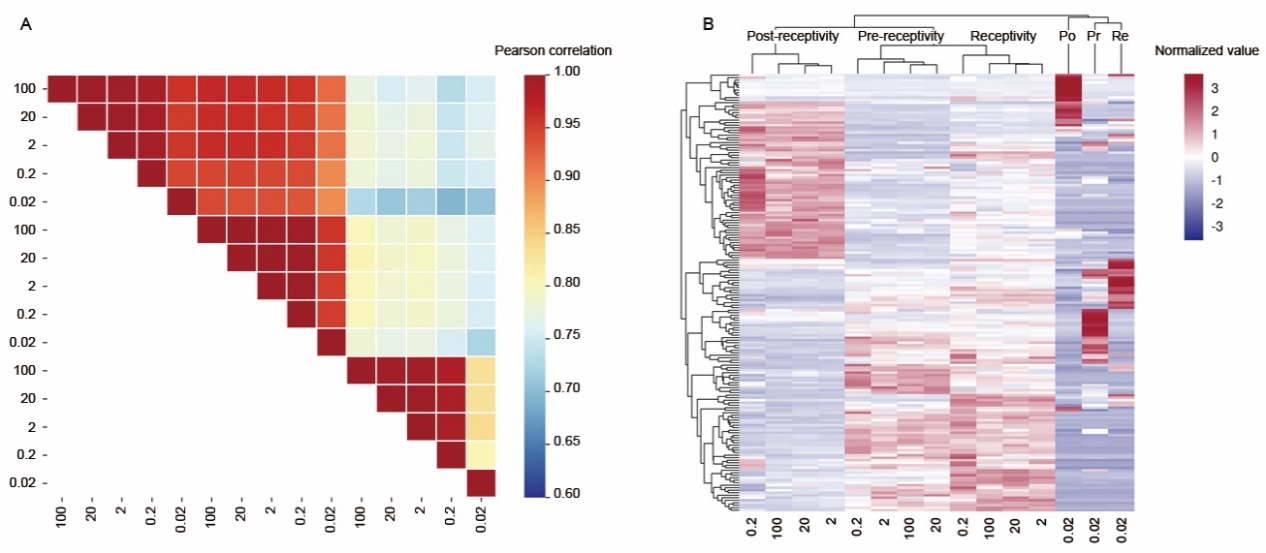

**Figure S1.** Analysis of stability and repeatability for the transcriptome sequencing with low amount of RNA. A. Pearson correlation between each library of different initial amounts of RNA. B. Hierarchical clustering of the top 200 differential expressed genes among three sequential uterine fluid samples.

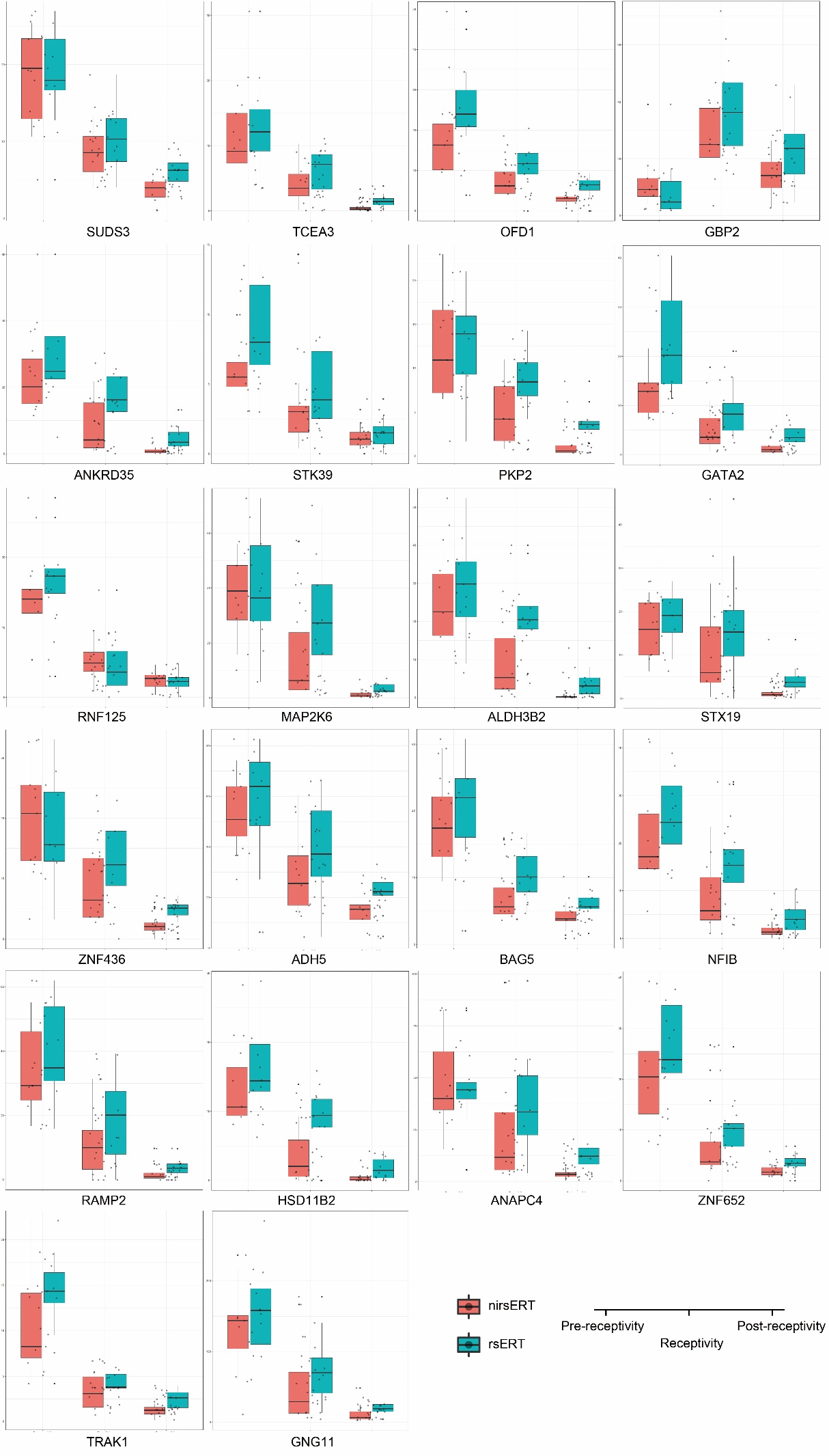

**Figure S2.** Expression pattern of 22 common markers between nirsERT and rsERT.

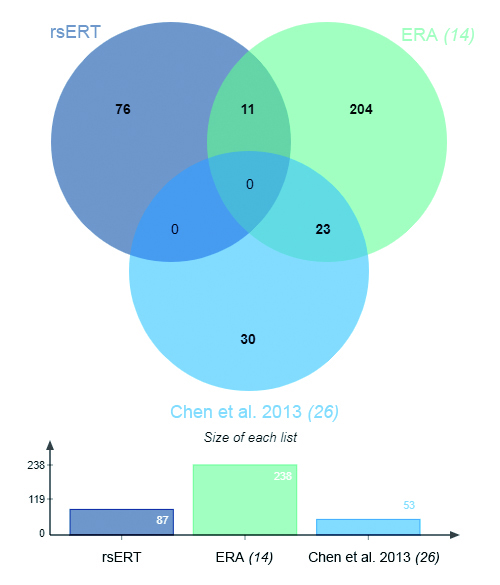

**Figure S3.** Venn diagram of predictive markers selected from three independent studies of endometrial receptivity. The Venn diagram is generated with jvenn (*2*).
